# Left ventricular mass and global wall thickness – prognostic utility and characterization of left ventricular hypertrophy

**DOI:** 10.1101/2022.10.16.22280561

**Authors:** Magnus Lundin, Einar Heiberg, David Nordlund, Tom Gyllenhammar, Katarina Steding-Ehrenborg, Henrik Engblom, Marcus Carlsson, Dan Atar, Jesper van der Pals, David Erlinge, Rasmus Borgquist, Ardavan Khoshnood, Ulf Ekelund, Jannike Nickander, Raquel Themudo, Sabrina Nordin, Rebecca Kozor, Anish N Bhuva, James C Moon, Eva Maret, Kenneth Caidahl, Andreas Sigfridsson, Peder Sörensson, Erik B Schelbert, Håkan Arheden, Martin Ugander

## Abstract

**AIMS:** Cardiovascular magnetic resonance (CMR) can accurately measure left ventricular (LV) mass, which is associated with morbidity and mortality. Several measures related to LV wall thickness exist, with uncertain relative prognostic strength. We hypothesized that prognosis can be used to select an optimal measure of wall thickness for characterizing LV hypertrophy.

**METHODS AND RESULTS:** Subjects having undergone CMR were studied (cardiac patients, n=2543; healthy volunteers, n=100). A new measure, global wall thickness (GT, GTI if indexed to body surface area), and related measures, including mass/volume and mass/volume^2/3^, were calculated. GT was accurately calculated from LV mass and LV end-diastolic volume. Among patients with follow-up (n=1575, median follow-up 5.4 years), the most predictive measure of death or hospitalization for heart failure was LV mass index (LVMI) (hazard ratio (HR)[95% confidence interval] 1.16[1.12–1.20], p<0.001), followed by GTI (HR 1.14[1.09–1.19], p<0.001). Among patients with normal mass, volume, systolic function and absence of scar (n=326, median follow-up 5.8 years), the most predictive measure was GT (HR 1.62[1.35–1.94], p<0.001). GT and LVMI could characterize patients as having a normal LV mass and wall thickness, or concentric remodeling, concentric hypertrophy, or eccentric hypertrophy, and the three abnormal groups had worse prognosis than the normal group (p<0.05 for all).

**CONCLUSIONS:** LV mass is highly prognostic when mass is elevated (advanced disease), but global wall thickness is easily and accurately calculated, and adds value and discrimination amongst those with normal mass (early disease). The combination of these two measures can be used to characterize hypertrophy.

## Background

Accurate characterization of left ventricular (LV) hypertrophy (LVH) is important since an increased LV mass (LVM) due to various forms of hypertrophy and remodeling is both a risk factor for cardiovascular disease and also modifiable.^1–3^ Echocardiographic measurements of end-diastolic wall thickness and cavity size lead to the standardized terminology of normal wall thickness and size, concentric hypertrophy, eccentric hypertrophy, and concentric remodeling.^4^ Cardiovascular magnetic resonance (CMR) provides additional accuracy^5^ and precision,^6^ via direct measurement of LV myocardial volumes and mass.

However, LVM alone does not allow for an assessment of wall thickness. There are several measures calculated from LVM and LV end-diastolic volume (LVEDV) that are related to wall thickness, such as mass:volume ratio (MVR=LVM/LVEDV) and concentricity^0.67^ (=LVM/LVEDV^2/3^). We hypothesized that a wall thickness-related measure could be used in combination with LVMI to characterize patients as having concentric remodeling (normal LVMI but high wall thickness), or as having hypertrophy (increased LVMI) that is either concentric (high wall thickness) or eccentric (low wall thickness). We further hypothesized that a measure such as global wall thickness (GT) could be easily calculated from LVM and LVEDV without the need for any special software, and that GT might have prognostic utility. This study sought to compare GT to other measures of LV hypertrophy by using prognostic association to evaluate which wall thickness-related measure would have greatest clinical relevance.

Therefore, the aims of the study were: (a) to use CMR to describe a new measure of global wall thickness (GT) that could be easily calculated, (b) to determine relative prognostic performance for LV mass, GT, and several other wall thickness-related measures, and (c) to classify LVH based on the measures with the highest prognostic associations.

## Methods

Cohorts (total *n*=2543) were selected from different cardiac disease states in order to address the respective aims. Figure 1 shows a schematic illustration of the composition of the cohorts and the intended use for each cohort.

**Figure 1.**
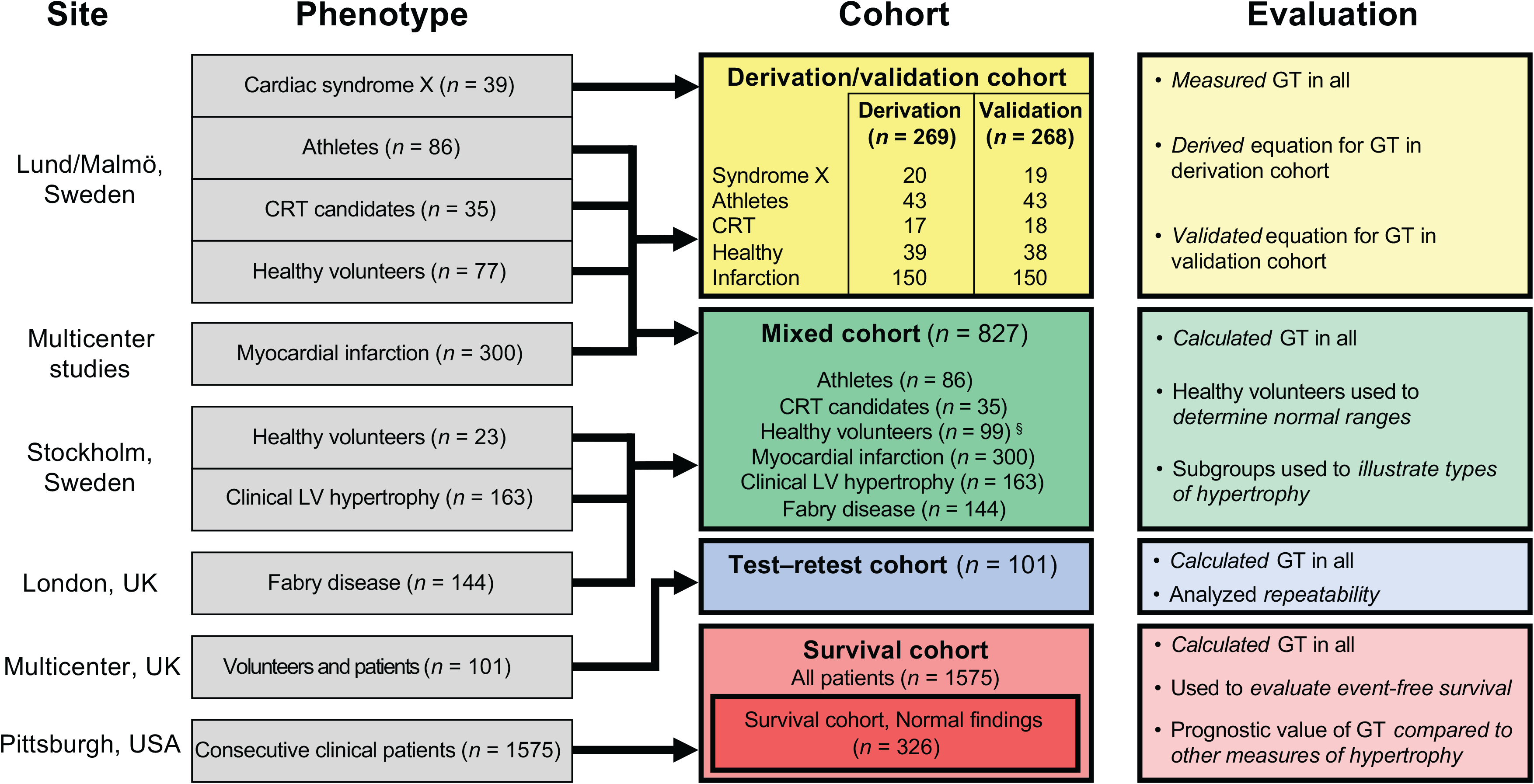
Schematic summary of the composition of the respective cohorts and what they were used to evaluate. CRT=cardiac resynchronization therapy, GT=global wall thickness; LV=left ventricular. ^§^ One of the healthy volunteers was an outlier and was excluded, see Methods.

All subjects provided written informed consent and were included following approval of the local human subject research ethics review board at the respective institutions. All subjects in this study underwent cine steady-state free precession (SSFP) imaging in a LV short-axis image stack for the analysis of LVEDV and LVM, excluding papillary muscles and trabeculations. BSA was calculated using the Mosteller method,^7^ and GT index (GTI) as GT/BSA.

### Derivation and validation cohort

In order to obtain and validate our new measure, we used a cohort (*n=*537) including healthy volunteers, athletes, patients with heart failure, recent myocardial infarction, and cardiac syndrome X. See Figure 1 and the Online supplementary data for details regarding the calculation, derivation and validation of GT.

Once derived, both GT and other measures related to hypertrophy such as mass:volume ratio, and concentricity^0.67 8^ were calculated for all subjects in order to compare their diagnostic and prognostic abilities. The remodeling index (RI) defined as (LVEDV)^1/3^/max wall thickness^9^ was calculated as (LVEDV)^1/3^/GT since maximum segmental wall thickness was not available.

### Survival cohort

Prognostic analysis was performed in a separate cohort (*n*=1575) of consecutive clinical patients investigated for cardiac disease. These patients underwent clinical CMR scans at 1.5 T (Siemens Magnetom Espree, Erlangen, Germany) between June 2010 and March 2016 at the University of Pittsburgh Medical Center (Pittsburgh, Pennsylvania, USA). The scan included cine SSFP imaging and late gadolinium enhancement (LGE) 10 minutes after a 0.2 mmol/kg dose of intravenous contrast agent (ProHance, Bracco Diagnostics, Stillwater, Minnesota, USA). This cohort has been examined regarding prognosis in several published studies^10^ but has never been studied with the objective of classifying hypertrophy. Patients with amyloidosis were excluded as they have a distinctive phenotype, and have prognostic features that markedly differ from other diseases. Exams that had previously been found to be of a sub-standard quality were also excluded.

All measures of LVH were examined regarding their prognostic value. A composite end-point was used, where the time from CMR exam to hospitalization for heart failure (HHF) or death from all causes was determined as previously described, and where HHF was confirmed by two cardiologists blinded to CMR data.^11^ Since both ventricular dilatation, low ejection fraction, and hypertrophy are important risk factors associated with heart failure,^1,12^ the analysis was performed both for the whole cohort as well as in the subgroup of patients with completely normal findings defined as LVEDVI, LVMI, and LV ejection fraction (LVEF) within the normal ranges, and absence of myocardial scar. The normal ranges used to select this subgroup were derived from the healthy volunteers described above.

### Test–retest cohort

In order to evaluate the repeatability of the old and new measures, a separate multicenter cohort (*n*=101) including different pathologies as well as healthy volunteers was used. The subjects were scanned at different magnetic field strengths at several locations in the United Kingdom between September 2010 and May 2019. These subjects were all scanned on two occasions, 96% were performed within one week and 79% on the same day. All analyses were undertaken by one expert observer.

### Mixed cohort

To obtain robust normal values, additional healthy volunteers (*n*=23, 39% female) underwent CMR at 1.5 T (Siemens Aera, Erlangen, Germany) at Karolinska University Hospital (Solna, Sweden) between April and August 2016 and were added to the healthy volunteers from the *derivation/validation cohort*, resulting in a total of 100 healthy volunteers. One of the female volunteers had an abnormal GTI four standard deviations above the mean GTI of the other female volunteers, in spite of having a LV mass and volume within the normal range. This outlier case was therefore not used for determining normal ranges. The resulting group of healthy volunteers (*n*=99, 35% female) was used to determine sex-specific 95% normal ranges for GT, GTI, LVEDVI, LVMI, and for the other measures listed above.

In order to illustrate the performance of the new measures for additional specific diagnoses, two additional groups of patients were included in the *mixed cohort*. Patients with prominent LVH (*n*=163, 37% female), were selected as having LVMI more than three standard deviations above the normal sex-specific mean calculated from the healthy volunteers, and underwent CMR at 1.5 T (Siemens Aera, Erlangen, Germany) at Karolinska University Hospital (Solna, Sweden) between October 2013 and November 2015. The second group was patients with genetically confirmed Fabry disease (*n*=144, 60% female), and were examined at 1.5 T (Siemens Avanto, Erlangen, Germany) at The Heart Hospital (London, United Kingdom) between May 2011 and July 2016. These groups were not included in any other cohort. Although the cohorts are inhomogeneous, they were included for different and well-defined purposes, as per Figure 1.

### Statistics

Calculations were performed using IBM SPSS Statistics (version 25, IBM Corporation, Armonk, New York, USA). Data are reported as mean±standard deviation (SD) or median [interquartile range] as appropriate. Normal ranges were defined as the range between the mean-1.96 SD and the mean+1.96 SD. To combine comparison of both sexes, measures were standardized to the sex-specific normal mean and reported as SD. The Mann–Whitney *U* test was used to compare non-normal distributions. Survival analysis was performed using SAS 9.4 (SAS Institute, Cary, North Carolina, USA). Cox regression was used, and univariable Cox regression χ^2^ values and hazard ratios (HR) were compared for different CMR parameters, standardized using the normal mean and SD per sex. The χ^2^ values were used to rank the different measures. The influence of different measures were also compared using multivariable Cox regression. Kaplan-Meier curves were plotted for the different types of hypertrophy and differences in prognosis were evaluated using the log-rank test. Test-retest variability was calculated as the relative Dahlberg error^13^ and the *F*-test was used to compare variances. Differences in prevalence were evaluated using the χ^2^-test. Statistical significance was defined as *p*<0.05.

## Results

The derivation/validation cohort (*n=*537) was used to derive and validate an equation for calculating GT in millimeters from LVM in grams and EDV in milliliters, as *GT* = 0.05 + 1.60 • *LVM* ^0.84^ • *LVEDV*^-0.49^ with excellent accuracy and precision, see the Online supplement. This equation can be used for any CMR results where only the values for LVM and LVEDV are available. Consequently, no special plugin or software is needed to calculate GT in clinical routine.

### Prognostic analysis

In the *survival cohort* (*n*=1575, 42% female, follow-up 5.4 [3.9–6.4] years), univariable Cox regression showed that, apart from age at CMR and presence of hypertension, the parameter with the highest prognostic value for hospitalization for heart failure (HHF) or death was LVMI (χ^2^ 66.7, *p<*0.001) followed by GTI (χ^2^ 37.3, *p<*0.001) and GT (χ^2^ 33.1, *p<*0.001). In multivariable analysis including LVMI and GTI, LVMI was associated with outcomes (*p*<0.001) and GTI was not (*p*=0.60), see Table 1.

**Table 1.**
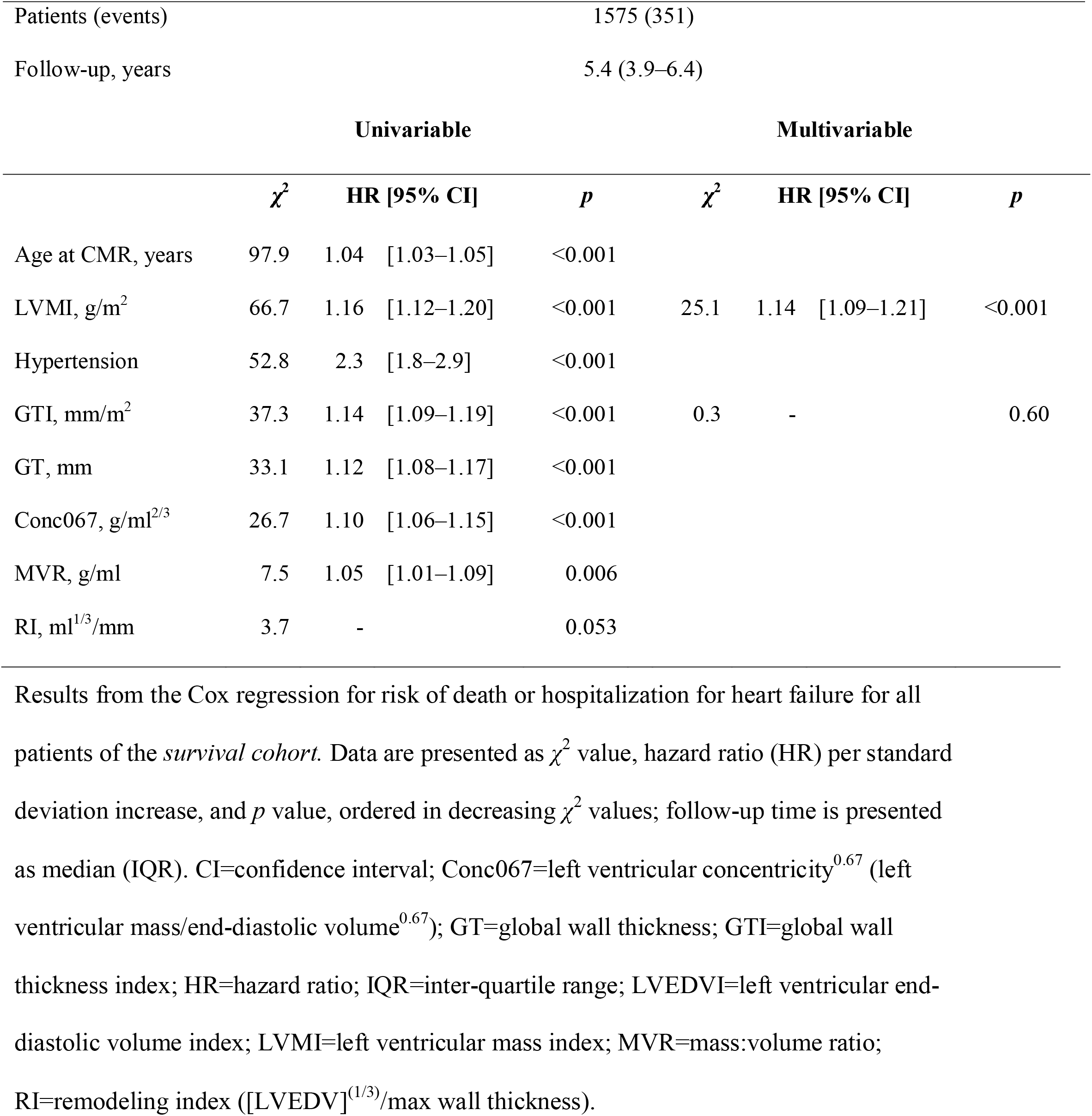
Prognostic results, all patients.

In the subgroup with normal findings (no LGE, and findings within the normal range per sex for LVEDVI, LVMI, and LVEF, *n*=326, 45% female, follow-up 5.8 [5.0–6.7] years), the parameters with the highest prognostic value for HHF or death were GT (χ^2^ 26.8, *p<*0.001), and concentricity^0.67^ (χ^2^ 26.6, *p<*0.001), and these were more prognostic than both hypertension (χ^2^ 23.5, *p<*0.001) and age at CMR (χ^2^ 14.5, *p*<0.001). In multivariable analysis including LVMI and GT, LVMI was not associated with outcomes (*p=*0.70) but GT was (*p=*0.01), see Table 2.

**Table 2.**
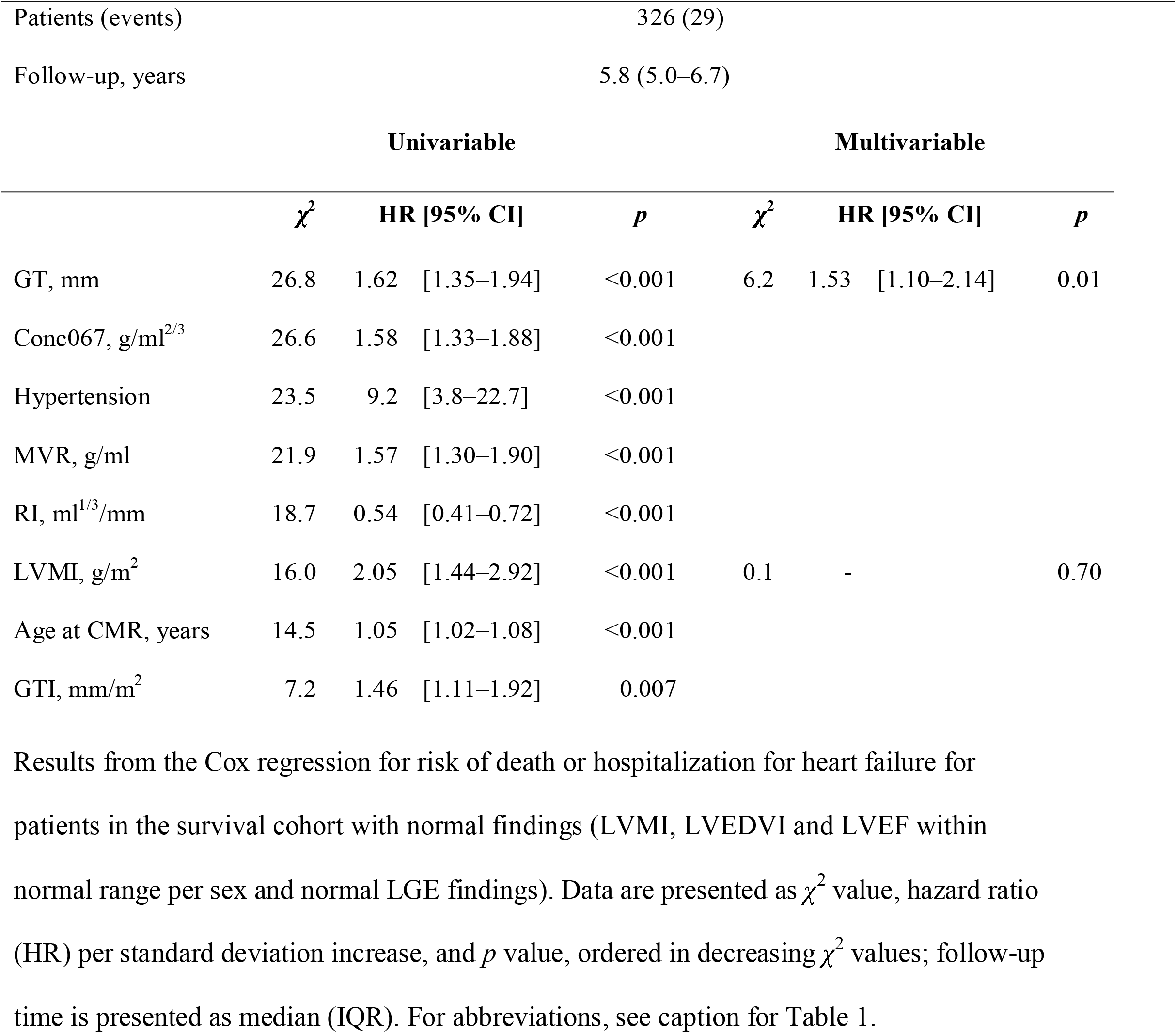
Prognostic results, patients with normal findings.

### Test–retest

In the test-retest cohort (*n*=101), the test-retest variability was lowest for GT (4.2%) and the highest variability was found for EDV and mass:volume ratio (6.1% and 6.2%, respectively, *p*<0.001 for both versus GT).

### Normal ranges

In the mixed cohort, normal calculated GT (based on healthy volunteers (*n=*99, 35% female), was 5.9±0.6 mm for females and 7.2±0.7 mm for males. This corresponds to a GT normal range of 4.8–7.1 mm for females and 5.8–8.5 mm for males. All patient groups, as well as the athletes, had a higher GT than healthy volunteers for both sexes (*p*<0.02 for all groups separately). Patient characteristics of the mixed cohort are presented in the Online supplement, Table S2.

When GT was corrected for body size (GTI), mean values were 3.4±0.4 mm/m^2^ for females and 3.6±0.4 mm/m^2^ for males, corresponding to normal ranges of 2.7–4.1 mm/m^2^ for females and 2.9–4.3 mm/m^2^ for males.

The athletes and all patient groups except for CRT candidates had higher GTI than healthy volunteers for both sexes (*p*<0.02 for all, except for CRT candidates). LVMI for the healthy volunteers was 50±7 g/m^2^ for females and 64±9 g/m^2^ for males and LVEDVI was 87±11 ml/m^2^ for females and 98±14 ml/m^2^ for males.

The mean LVM among healthy volunteers was 2.4 SD higher in males compared to females, whereas mean GT among healthy volunteers was 2.0 SD higher for males compared to females.

### Classification of left ventricular hypertrophy subtypes

As illustrated in the flow chart in Figure 2, the combination of GT and LVMI can be used to characterize patients as being normal (normal GT, normal LVMI), or having concentric remodeling (high GT, normal LVMI), eccentric hypertrophy (high LVMI, normal GT), or concentric hypertrophy (high LVMI, high GT).

**Figure 2.**
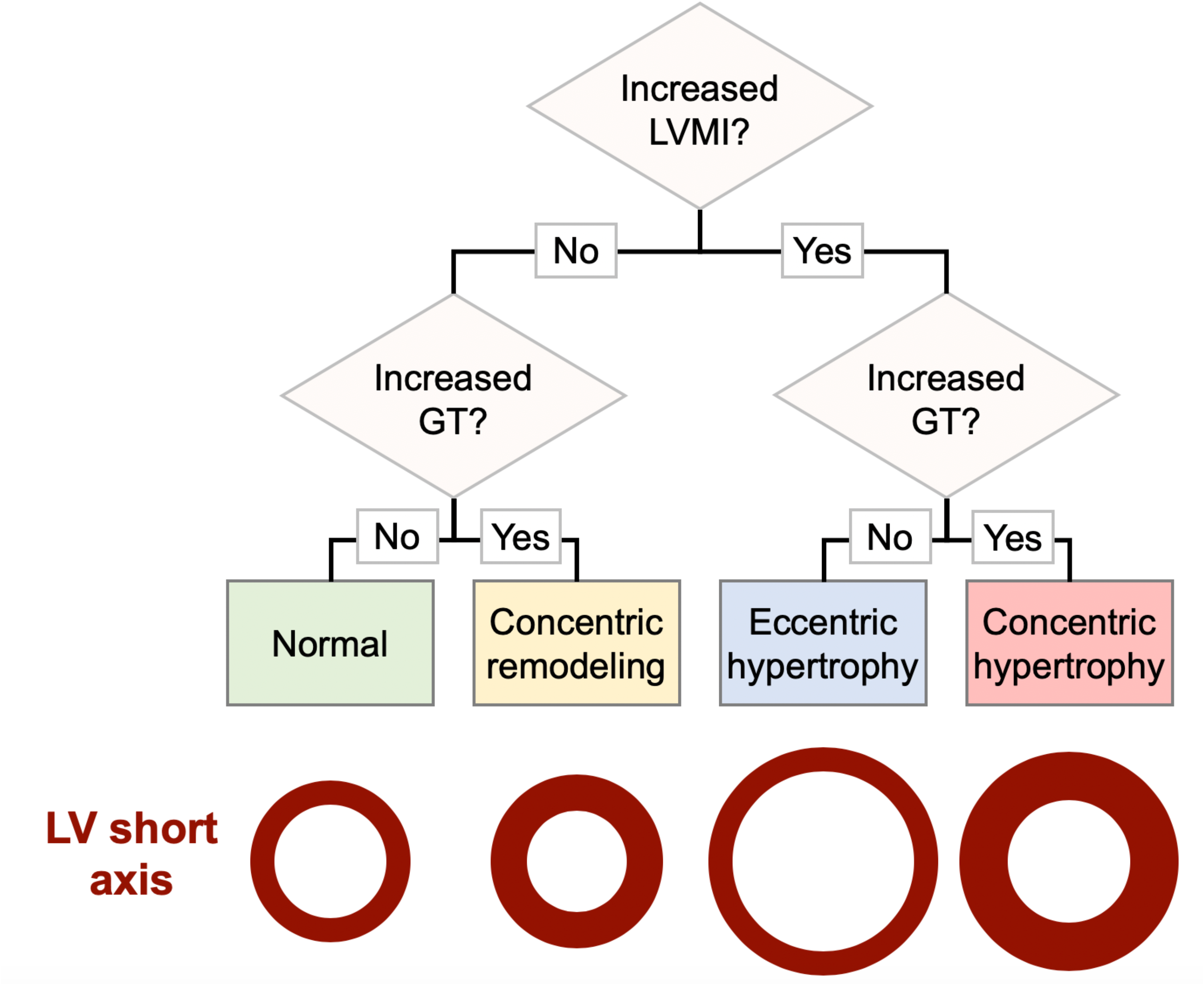
Proposed flow chart for characterizing different types of LV hypertrophy and remodeling. The bottom of the image includes a schematic illustration of a typical LV short axis slice for each classification outcome. BSA=body surface area; GT=global wall thickness indexed to body surface area (BSA); LV=left ventricular; LVMI=LV mass indexed to BSA.

In the survival cohort (*n=*1575), the combination of GT and LVMI classified 1133 patients (72%) as being normal, 189 patients (12%) as having concentric remodeling, 89 patients (6%) as having eccentric hypertrophy, and 164 patients (10%) as having concentric hypertrophy.

Patients with concentric remodeling had worse prognosis, for death or HHF, than patients classified as normal (*p=*0.004). Both the group of patients with concentric hypertrophy and the group with eccentric hypertrophy had worse prognosis than the normal group (*p*<0.0001 for both), and since concentric and eccentric hypertrophy did not differ from one another regarding prognosis (*p*=0.66), they were also analyzed as one hypertrophy group (*n=*253, 16% of all patients). These patients with hypertrophy had worse prognosis than both patients classified as normal (*p*<0.0001) and patients with concentric remodeling (*p*=0.003), see Figure 3. Patient characteristics for the survival cohort are shown in the Online supplement Tables S3 and S4.

**Figure 3.**
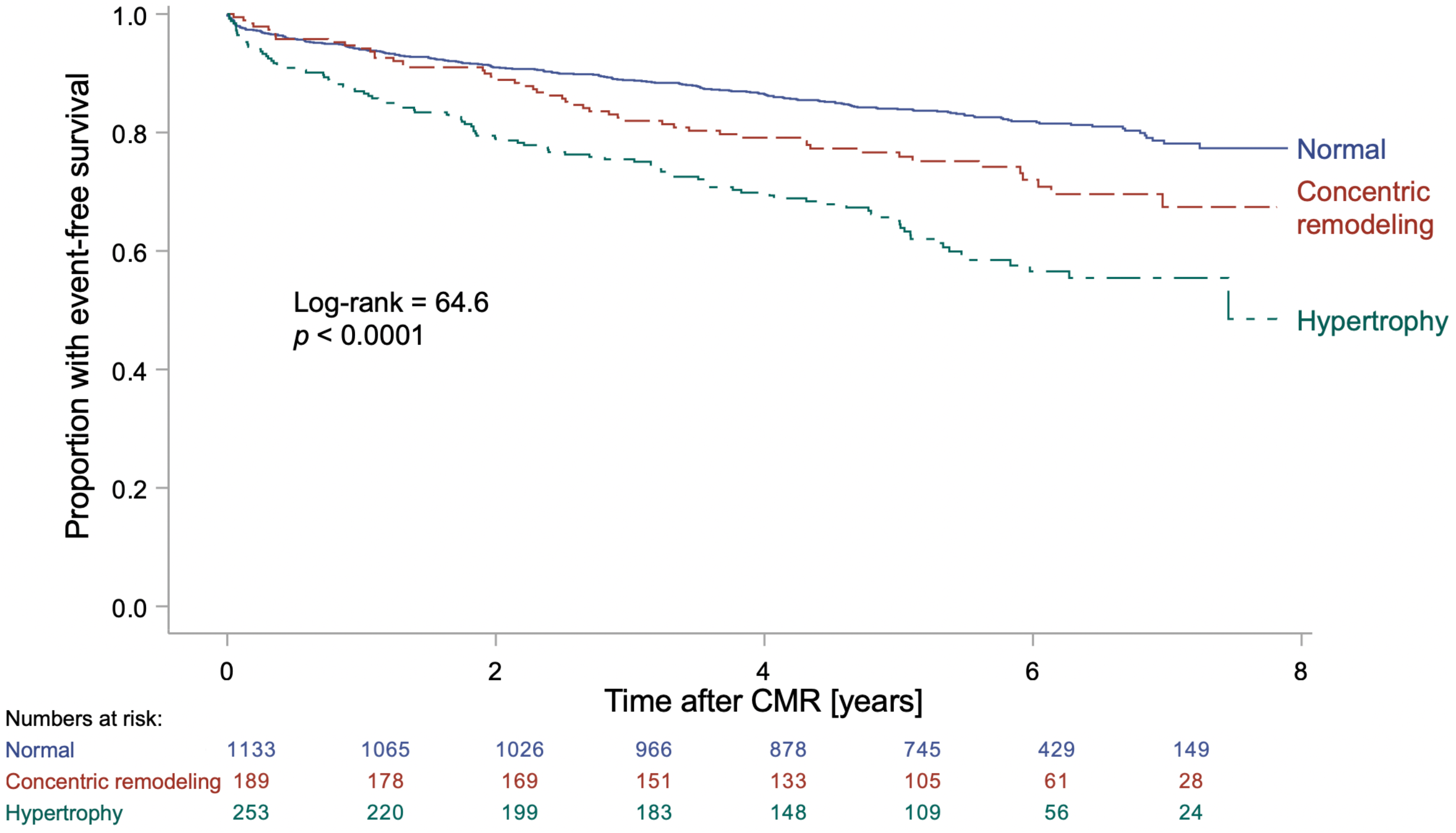
Kaplan-Meier curves for the survival cohort. Kaplan-Meier curve for the consecutive clinical patients of the *survival cohort* (*n*=1575, follow-up 5.4 [3.9–6.4] years) classified as having either hypertrophy (increased LVMI regardless of GT as a combined group); concentric remodeling (normal LVMI, increased GT); or being classified as normal (normal LVMI, normal GT). Event-free survival was defined as absence of the combined endpoint of death or hospitalization for heart failure. The patients with hypertrophy had worse prognosis compared to both the concentric remodeling (*p*=0.003) and the normal group (*p*<0.0001). Patients with concentric remodeling had worse prognosis compared to the normal group (*p*=0.004). An increase in LVMI or GT was based on the 95% upper limit of normal calculated from the healthy volunteers for females and males respectively. CMR=cardiovascular magnetic resonance; GT=global wall thickness; LVMI=left ventricular mass indexed to body surface area.

When evaluating the classification of the patient groups in the mixed cohorts, the majority of patients with Fabry disease were classified as having concentric remodeling, albeit with a large variability due to the variability in the manifestation of the disease. The majority of patients selected for having prominent LVH were classified as having concentric hypertrophy, most CRT patients were classified as having eccentric hypertrophy, and the medians for all other groups were in the normal range, see Figure 4.

**Figure 4.**
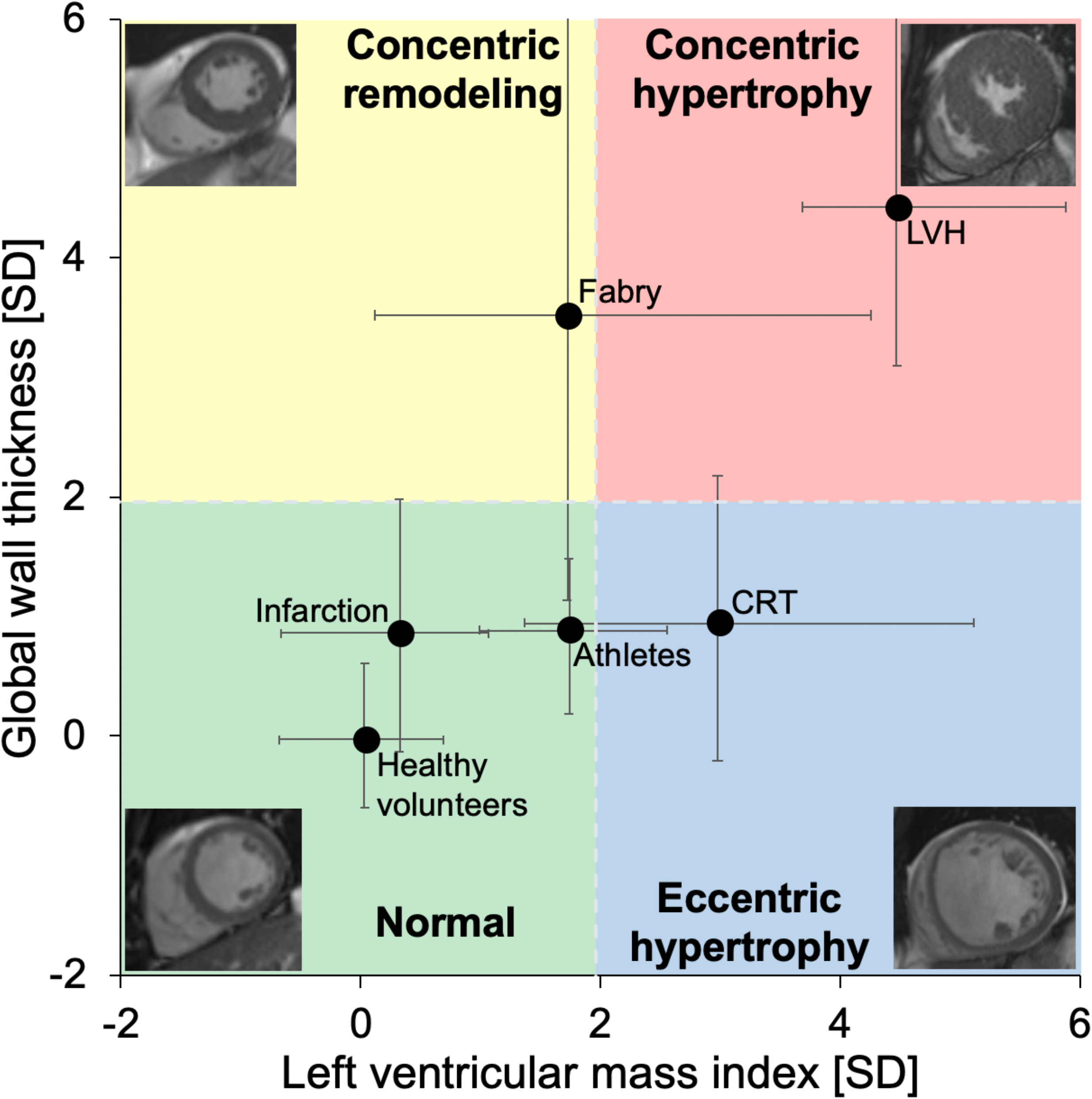
Characterization of left ventricular hypertrophy using wall thickness and mass. Global wall thickness (GT) plotted versus left ventricular mass index (LVMI) for the *mixed cohort*, who were not used in the prognostic analysis. The solid circles show the median and the whiskers show the interquartile range. Both GT and LVMI have been standardized to standard deviations (SD) from the sex-specific mean of the healthy volunteers. The colored fields show the proposed classification of hypertrophy based on LVMI and GT. The gray dashed lines indicate the upper limit of normal (+1.96 SD) for both GT and LVMI. The mixed cohort consist of healthy volunteers, endurance athletes, cardiac resynchronization therapy (CRT) candidates, patients with recent acute ST-elevation myocardial infarction (Infarction), patients with Fabry disease (Fabry), and patients with at least moderate left ventricular hypertrophy (LVH). Four examples of the proposed classification of hypertrophy are shown in the four corners. CMR=cardiovascular magnetic resonance; CRT=cardiac resynchronization therapy; LVH=left ventricular hypertrophy; LVMI=left ventricular mass index; SD=standard deviations; GT=global wall thickness.

## Discussion

This study shows that global wall thickness (GT) can be calculated from LV mass and end diastolic volume using a formula without the need for any dedicated plugin or software. The prognostic analysis shows that LVMI was the most prognostic measure of hypertrophy regarding death or hospitalization for heart failure in the survival cohort as a whole, followed by GTI. However, among patients with normal findings, GT was the most prognostic measure, and it was more prognostic than both age and hypertension. The hazard ratios for GT and GTI were larger than 1 in the univariable analysis for all examined groups indicating that a thicker wall is always a negative prognostic factor.

These findings illustrate the prognostic difference between an increased LVMI seen in advanced hypertrophy, and an increase in GT which may precede overt LV hypertrophy. Among the patients in the *survival cohort* with normal findings, GT had a higher prognostic association than both the other hypertrophy measures, as well as hypertension and age. GT can therefore be used to identify concentric remodeling in patients with otherwise normal findings and monitor these changes over time. The findings indicate that GT has a high sensitivity for detecting small changes in left ventricular wall thickness and that these small changes carry prognostic information, although the mechanism on a cellular level for this remains unclear. Furthermore, it is possible to generate an accurate and precise measure of GT from whole heart CMR measurement of LV mass and volume. The repeatability was better for GT compared to other measures, and GT was found to be less sex-dependent than LVM.

Thus, using GT together with LVMI enables characterization of different types of LV hypertrophy: normal configuration, concentric remodeling, eccentric hypertrophy, or concentric hypertrophy. This classification requires only two measures (GT and LVMI), and the results can be visualized in a two-dimensional diagram, see Figure 4. There was no difference in outcomes between patients with eccentric and concentric hypertrophy in the survival cohort, and this is in line with the finding that LVMI was the predominant risk factor in the whole survival cohort. The distinction between eccentric and concentric hypertrophy could nevertheless imply different etiologies, necessitating different treatments.

Hypertrophy has previously been characterized using LV mass and relative wall thickness measured using echocardiography^4^ and a previous study found that the development of an abnormal LV mass and/or abnormal relative wall thickness was associated with an increased risk for cardiovascular disease.^14^ Since echocardiography only measures LV wall thickness in one or two locations, this method is inherently more prone to errors compared to the proposed method. However, since GT reflects the whole LV, it will not detect areas with asymmetric hypertrophy, but such areas should be easy to identify in CMR images. Dedicated software could also be used to measure the wall thickness per slice and/or segment, but such results could not be obtained using a simple formula such as for GT.

Our study illustrates how mass and volume are geometrically related and determine wall thickness, and how both are associated with events. A previous study found that LVMI and LVEDVI were both associated with heart failure events.^15^ While this is in agreement with our results, the current study clarifies how mass and volume together determine wall thickness and how both mass and wall thickness are different components of hypertrophy. Interestingly, mass and wall thickness have been shown to have distinctly different manifestations in the ECG^16^.

LV mass measured by CMR has been used to calculate measures such as mass:volume ratio^17^ and concentricity^0.67^,^8,18^ where the latter classification method requires three measures (concentricity^0.67^, LV mass, and LVEDV) for characterizing hypertrophy. By comparison, the proposed classification method is more simple since it only requires LVMI and GT (calculated from LVM and LVEDV). Concentricity^0.67^ performed well in the prognostic analysis, and the way that it is calculated is justified since an object that is twice as large in every direction will have a volume that is eight (2^3^) times larger, whereas the surrounding shell will only be four (2^2^) times as large. Indeed, calculating concentricity^0.67^ is somewhat similar to calculating GT (LVM vs LVM^0.84^, and LVEDV^-0.67^ vs LVEDV^-0.49^). Notably, GT has an advantage over Concentricity^0.67^ since GT provides the average wall thickness in mm, which is an intuitive measure.

GT was increased above the upper limit of normal in 14% of the patients with normal mass in the survival cohort. Previous studies using echocardiography and relative wall thickness to define concentric remodeling have found the prevalence of concentric remodeling to be 19%^4^ and 16%^19^ among hypertensive patients with normal LV mass, which is effectively similar to our results for GT.

## Limitations

The expression used to calculate GT uses both LVM and LVEDV derived from cine images. Separate data for papillary muscles and trabeculations was not available in the present study. These structures were not included in LVM and were included in the blood volume.

Therefore, the derived equation for GT is only applicable to results from exams where delineations are performed in a similar fashion. As with all cohort studies, the *prognostic cohort* is subject to some selection bias, but nevertheless reflects clinical practice and therefore remains inherently worthy of study. Only measures related to hypertrophy, as well as age, and no measures of systolic function were included in the prognostic analysis since the objective was to find a measure related to hypertrophy that is complementary to LVM, and not to identify the most prognostic measure overall.

It has been found that in healthy volunteers with blood pressure ≤140/90 mmHg, both mass and volume decrease with increasing age.^20^ However, calculations based on data from that publication show that there was negligible change in GT with age based on GT calculated from different age group means in that study [data not shown]. Thus, normal values for GT regardless of age are appropriate.

Only linear regression was used in the prognostic analysis. The finding that the hazard ratios for GT and GTI were larger than 1 for all examined groups does not, *per se*, indicate a relationship that is linear, but it indicates at least that a thicker wall implies a worse prognosis overall. Multiple linear regression was performed for LVMI and GT(I) where the latter is dependent on the first (correlated with an R^2^ of roughly 0.5). Nevertheless, for the patients with normal findings, the χ^2^ was much higher for GT in both the univariable and the multivariable analysis indicating a superior prognostic performance. The aim of the study was specifically to compare the relative prognostic strength of measures related to hypertrophy. Therefore, ejection fraction, scar burden and other measures were not included in the prognostic analysis.

The present study includes exams performed at several different centers, on several different platforms, and analyzed by different observers, which can contribute to variation in the results. However, CMR measurement of LVM has a measurement precision that vastly exceeds LVM measurements by 2D echocardiography,^21^ and thus variability in measurement of LVM and EDV would not be expected to have a sizeable impact on our results in comparison to the variability of echocardiographic approaches. Also, the test-retest repeatability was better for GT compared to LVM and LVEDV.

## Conclusions

LV mass (LVMI) is the most prognostic measure for death or hospitalization for heart failure among patients investigated with CMR for cardiac disease, whereas the global wall thickness (GT, calculated simply from LV mass and end-diastolic volume), is the most prognostic measure in patients with normal CMR findings. This suggests that an optimal measurement of LV hypertrophy is a combination of LVMI (advanced disease) and GT (early disease).

## Data Availability

All data produced in the present study are available upon reasonable request to the authors.

## Supplementary data

The Online supplementary data contains details regarding the calculation, derivation and validation of the new measure global wall thickness (GT) as well as characteristics of the derivation/validation cohort, the mixed cohort, and the survival cohort.

## References

1. Levy D, Garrison RJ, Savage DD, Kannel WB, Castelli WP. Prognostic implications of echocardiographically determined left ventricular mass in the Framingham Heart Study. N Engl J Med 1990;322:1561–1566.

2. Gupta S, Berry JD, Ayers CR, Peshock RM, Khera A, Lemos JA De, Patel PC, Markham DW, Drazner MH. Left ventricular hypertrophy, aortic wall thickness, and lifetime predicted risk of cardiovascular disease: The Dallas heart study. JACC Cardiovasc Imaging Elsevier Inc.; 2010;3:605–613.

3. Verdecchia P, Schillaci G, Borgioni C, Ciucci A, Battistelli M, Bartoccini C, Santucci A, Santucci C, Reboldi G, Porcellati C. Adverse prognostic significance of concentric remodeling of the left ventricle in hypertensive patients with normal left ventricular mass. J Am Coll Cardiol 1995;25:871–878.

4. Ganau A, Devereux RB, Roman MJ, Simone G de, Pickering TG, Saba PS, Vargiu P, Simongini I, Laragh JH. Patterns of left ventricular hypertrophy and geometric remodeling in essential hypertension. J Am Coll Cardiol 1992;19:1550–1558.

5. Caputo GR, Tscholakoff D, Sechtem U, Higgins CB. Measurement of canine left ventricular mass by using MR imaging. Am J Roentgenol 1987;148:33–38.

6. Grothues F, Smith GC, Moon JCC, Bellenger NG, Collins P, Klein HU, Pennell DJ. Comparison of interstudy reproducibility of cardiovascular magnetic resonance with two-dimensional echocardiography in normal subjects and in patients with heart failure or left ventricular hypertrophy. Am J Cardiol 2002;90:29–34.

7. Mosteller RD. Simplified calculation of body-surface area. N Engl J Med 1987;317:1098.

8. Khouri MG, Peshock RM, Ayers CR, Lemos JA De, Drazner MH. A 4-tiered classification of left ventricular hypertrophy based on Left ventricular geometry the dallas Heart study. Circ Cardiovasc Imaging 2010;3:164–171.

9. Goh VJ, L. TT, Bryant J, Wong JI, Su B, Lee CH, Pua CJ, Sim CPY, Ang B, Aw TC, Cook SA, Chin CWL. Novel index of maladaptive myocardial remodeling in hypertension. Circ Cardiovasc Imaging 2017;10.

10. Treibel TA, Fridman Y, Bering P, Sayeed A, Maanja M, Frojdh F, Niklasson L, Olausson E, Wong TC, Kellman P, Miller CA, Moon JC, Ugander M, Schelbert EB. Extracellular Volume Associates With Outcomes More Strongly Than Native or Post-Contrast Myocardial T1. JACC Cardiovasc Imaging 2020;13:44–54.

11. Schelbert EB, Piehler KM, Zareba KM, Moon JC, Ugander M, Messroghli DR, Valeti US, Chang CCH, Shroff SG, Diez J, Miller CA, Schmitt M, Kellman P, Butler J, Gheorghiade M, Wong TC. Myocardial fibrosis quantified by extracellular volume is associated with subsequent hospitalization for heart failure, death, or both across the spectrum of ejection fraction and heart failure stage. J Am Heart Assoc 2015;4.

12. Vasan RS, Larson MG, Benjamin EJ, Evans JC, Levy D. Left ventricular dilatation and the risk of congestive heart failure in people without myocardial infarction. N Engl J Med 1997;336:1350–1355.

13. Kim H-Y. Statistical notes for clinical researchers: Evaluation of measurement error 2: Dahlberg’s error, Bland-Altman method, and Kappa coefficient. Restor Dent Endod 2013;38:182.

14. Lieb W, Gona P, Larson MG, Aragam J, Zile MR, Cheng S, Benjamin EJ, Vasan RS. The natural history of left ventricular geometry in the community: Clinical correlates and prognostic significance of change in LV geometric pattern. JACC Cardiovasc Imaging 2014;7:870–878.

15. Bluemke DA, Kronmal RA, Lima JAC, Liu K, Olson J, Burke GL, Folsom AR. The Relationship of Left Ventricular Mass and Geometry to Incident Cardiovascular Events. The MESA (Multi-Ethnic Study of Atherosclerosis) Study. J Am Coll Cardiol American College of Cardiology Foundation; 2008;52:2148–2155.

16. Maanja M, Schlegel TT, Kozor R, Lundin M, Wieslander B, Wong TC, Schelbert EB, Ugander M. The electrical determinants of increased wall thickness and mass in left ventricular hypertrophy. J Electrocardiol Elsevier Inc.; 2020;58:80–86.

17. Rosen BD, Edvardsen T, Lai S, Castillo E, Pan L, Jerosch-Herold M, Sinha S, Kronmal R, Arnett D, Crouse JR, Heckbert SR, Bluemke DA, Lima JAC. Left ventricular concentric remodeling is associated with decreased global and regional systolic function: the Multi-Ethnic Study of Atherosclerosis. Circulation 2005;112:984–991.

18. Bang CN, Gerdts E, Aurigemma GP, Boman K, Dahlöf B, Roman MJ, Køber L, Wachtell K, Devereux RB. Systolic left ventricular function according to left ventricular concentricity and dilatation in hypertensive patients: The Losartan Intervention for Endpoint reduction in hypertension study. J Hypertens 2013;31:2060–2068.

19. Verdecchia P, Porcellati C, Zampi I, Schillaci G, Gatteschi C, Battistelli M, Bartoccini C, Borgioni C, Ciucci A. Asymmetric left ventricular remodeling due to isolated septal thickening in patients with systemic hypertension and normal left ventricular masses. Am J Cardiol 1994;73:247–252.

20. Cain PA, Ahl R, Hedstrom E, Ugander M, Allansdotter-johnsson A, Friberg P, Arheden H. Age and gender specific normal values of left ventricular mass, volume and function for gradient echo magnetic resonance imaging: a cross sectional study. BMC Med Imaging 2009;9:1–10.

21. Bellenger NG, Davies LC, Francis JM, Coats AJ, Pennell DJ. Reduction in sample size for studies of remodeling in heart failure by the use of cardiovascular magnetic resonance. J Cardiovasc Magn Reson 2000;2:271–278.

