## Supplementary data for "Left ventricular mass and global wall thickness – prognostic utility and characterization of left ventricular hypertrophy"

### Online supplementary data

***Derivation and validation of global wall thickness.*** To derive and validate the new measure global wall thickness (GT), a large representative cohort of health and disease (*n=*537) including healthy volunteers, athletes, patients with heart failure, recent infarction, cardiac syndrome X was used.

Healthy volunteers had no heart disease, no hypertension, no present or previous systemic or cardiovascular disease, were non-smokers, and did not use any medications with known cardiovascular effects, they were scanned between May 2001 and May 2009. Athletes were elite endurance athletes at national level competition in either soccer, European handball, swimming, or triathlon, they were scanned between December 2005 and March 2008. Heart failure patients were candidates for cardiac resynchronization therapy (CRT) and were scanned between December 2010 and November 2014. The patients with myocardial infarction were part of multicenter trials of acute myocardial infarction and were scanned between August 2011 and July 2015. Cardiac syndrome X were scanned between November 2011 and February 2016.

The athletes, the healthy volunteers and the patients with heart failure and cardiac syndrome X were all examined at Skåne University Hospital, Lund or Malmö, Sweden, and were scanned at 1.5 T (Siemens Aera, Erlangen, Germany, or Philips Intera, Best, the Netherlands). For all groups, the epicardial and endocardial borders were delineated manually and the end-diastolic and end-systolic volumes as well as LVM (mean of end-systolic and end-diastolic mass) were calculated using the freely available software Segment (http://segment.heiberg.se) (19). To evaluate systolic function in terms of ejection fraction and end-systolic volumes as well as comparing end-systolic and end-diastolic LVM was not the aim of the current study, which specifically focused on comparative prognostic measures related to LV mass and wall thickness.

An in-house developed plug-in for the software Segment was used to supply the measurements used to derive and validate GT. A schematic illustration of the method for measuring GT is shown in Figure S1. The plug-in automatically measured the distance between the endocardial and epicardial borders at 24 evenly distributed positions around the circumference of each short-axis slice in end-diastole. Regions with a wall thickness of less than 2 mm in the left ventricular outflow tract or apex were excluded. The mean wall thickness of each short-axis slice was multiplied by the midmural circumference of the respective slice, and the resulting sum for all slices was divided by the sum of the circumferences of all slices to yield the GT, thereby weighting wall thickness by slice size.

In order to be able to *calculate* the GT from known parameters without the use of dedicated software, a simple equation was proposed. Since GT geometrically can only depend on cardiac mass and volume, GT was mathematically expressed as relating to LVM and LVED according to the following equation:

*GT* = *A* + *B* • *LVM^X^* • *LVEDV^Y^*	(Eq. 1),

where GT is mean left ventricular end-diastolic global wall thickness in millimeters, LVM is left ventricular mass in grams, and LVEDV is left ventricular end-diastolic volume in milliliters, and *A*, *B*, *X*, and *Y* are constants to be estimated. The included groups were split into a derivation and validation cohort matched for sex and diagnosis. Matlab (R2016, Mathworks, Natick, Massachusetts, USA) was used to identify best fits for *A*, *B*, *X* and *Y* (Eq. 1). Fit performance was estimated by least squares of data against all possible combinations of *A*, *B*, *X* , and *Y* in the derivation cohort, with performance evaluated subsequently in the validation cohort.

#### Derivation, validation and normal ranges for GT

The patient characteristics of the derivation and validation cohorts are shown in Table S1. Using the derivation subset (*n*=269) of the derivation/validation cohort, the optimized equation for the calculated global wall thickness (GT) was found to be:

*GT =* 0.05 + 1.60 *• LVM*^0.84^ *• LVEDV*^-0.49^	(Eq. 2).

Where GT is the global wall thickness in mm, LVM is left ventricular mass in grams, and LVEDV is left ventricular end-diastolic volume in milliliters. For the derivation subset, the model had an expectedly high correlation (*R*^2^=0.95, *p* < 0.001) with no bias (0.00±0.24 mm), see Figure S2, upper panels. When applied to the separate *validation* *subset* of the cohort (*n=*268) model performance was preserved (*R*^2^=0.95, *p* < 0.001, bias 0.01±0.23 mm), see Figure S2, lower panels. The wide range of values for LV mass and volumes in the various patient groups of the derivations and validation cohorts (see Table S1) imply that the derived equation should be valid for a wide range of combinations of LV size, mass and wall thickness.

Normal calculated GT (based on healthy volunteers (*n=*99, 35% female), was 5.9±0.6 mm for females and 7.2±0.7 mm for males. This corresponds to a GT normal range of 4.8–7.1 mm for females and 5.8–8.5 mm for males. All patient groups, as well as the athletes, had a higher GT than healthy volunteers for both sexes (*p* < 0.02 for all groups separately).

When GT was corrected for body size (GTI), males and females were much closer together 3.4±0.4 mm/m^2^ for females and 3.6±0.4 mm/m^2^ for males, corresponding to normal ranges of 2.7–4.1 mm/m^2^ for females and 2.9–4.3 mm/m^2^ for males. LVMI for the healthy volunteers was 50±7 g/m^2^ for females and 64±9 g/m^2^ for males and LVEDVI was 87±11 ml/m^2^ for females and 98±14 ml/m^2^ for males.

Figures

**Figure S1.** Schematic illustration of how mean left ventricular (LV) end-diastolic global wall thickness (GT) was measured using a LV short-axis image stack. The distance between the endocardial and epicardial borders at end-diastole was measured at 24 evenly distributed positions (shown as dashed black lines) around the circumference of all short-axis slices of a full LV short-axis stack from base to apex. Basal sections with a wall thickness of less than 2 mm in the LV outflow tract and the apex were excluded. The mean thicknesses for each individual short-axis slice was multiplied by the midmural circumference of that slice (pink circle), these were summed and then divided by the sum of the midmural circumference for all slices to yield the GT. GT=global wall thickness; LV=left ventricular.

**Figure S2.** Plots of the calculated vs. measured global wall thickness (GT) in mm. GT was measured using the method illustrated in Figure S1, and was estimated using a derived formula, see the Online appendix for details. Top left: Correlation plot for the derivation subset (n=269), *R*^2^=0.95, bias 0.00±0.24 mm, identity line shown dashed. Top right: Bland–Altman plot for the derivation subset of the cohort. Solid line shows mean difference and dashed lines show +/- 1.96 standard deviations. Bottom left: Correlation plot for the separate validation subset (*n=*268), *R*^2^=0.95, *p* < 0.001, bias 0.01±0.23 mm, identity line shown dashed. Bottom right: Bland–Altman plot for the validation subset of the cohort. Solid line shows mean difference and dashed lines show +/- 1.96 standard deviations. GT=global wall thickness.

### Tables

**Table S1a. Characteristics for the female subjects of *derivation/validation cohort*.**

| ***Females*** | **Healthy volunteers** | | **Athletes** | | **CRT** | | **Infarction** | | **Cardiac**  **syndrome X** | |
| --- | --- | --- | --- | --- | --- | --- | --- | --- | --- | --- |
| Number, *n* | 27 |  | 41 |  | 7 |  | 64 |  | 18 |  |
| Age, years | 36 | (26–60) | 22 | (19–26)* | 66 | (63–74)* | 69.5 | (60–75)* | 69.5 | (57–74)* |
| Length, cm | 169 | (165–170) | 170 | (167–174) | 163 | (159–174) | 165 | (160–171) | 163 | (160–168)* |
| Weight, kg | 66 | (59–70) | 64 | (60–70) | 67 | (59–80) | 73 | (66–80)* | 71 | (64–79) |
| BSA, m^2^ | 1.8 | (1.7–1.8) | 1.7 | (1.7–1.8) | 1.8 | (1.6–1.9) | 1.8 | (1.7–1.9)* | 1.8 | (1.7–1.9) |
| BMI, kg/m^2^ | 23 | (21–25) | 22 | (21–23) | 25 | (22–30) | 26 | (24–28)* | 26 | (25–28)* |
| LVEF, % | 61 | (59–63) | 57 | (56–59)* | 26 | (18–36)* | 52 | (45–58)* | 68 | (66–70)* |
| LVEDV, ml | 156 | (131–169) | 188 | (175–204)* | 300 | (196–392)* | 131 | (114–170)* | 126 | (118–144)* |
| LVESV, ml | 61 | (50–72) | 82 | (73–88)* | 215 | (124–321)* | 65 | (49–81) | 39 | (35–47)* |
| LVSV, ml | 91 | (83–104) | 107 | (99–116)* | 72 | (56–106) | 69 | (57–83)* | 87 | (78–98) |
| LVM, g | 88 | (74–94) | 111 | (105–117)* | 167 | (107–210)* | 98 | (86–113)* | 90 | (81–98) |
| LVEDVI, ml/m^2^ | 90 | (77–97) | 109 | (99–115)* | 181 | (117–236)* | 75 | (64–88)* | 70 | (66–83)* |
| LVESVI, ml/m^2^ | 28 | (36–39) | 41 | (47–49)* | 67 | (120–193)* | 28 | (36–45) | 20 | (22–27)* |
| LVSVI, ml/m^2^ | 53 | (48–59) | 62 | (58–65)* | 43 | (32–58) | 40 | (32–46)* | 47 | (45–54) |
| LVMI, ml/m^2^ | 51 | (42–54) | 64 | (57–69)* | 93 | (62–115)* | 55 | (48–62)* | 51 | (46–55) |

* denotes *p* < 0.05 compared to the healthy volunteers. **Table S1b. Characteristics for the male subjects of *derivation/validation cohort*.**

| ***Males*** | **Healthy volunteers** | | **Athletes** | | **CRT** | | **Infarction** | | **Cardiac**  **syndrome X** | | | |
| --- | --- | --- | --- | --- | --- | --- | --- | --- | --- | --- | --- | --- |
| Number, *n* | 50 |  | 45 |  | 28 |  | 236 |  | | 21 |  | |
| Age, years | 34 | (27–51) | 26 | (21–33)* | 69 | (65–74)* | 59 | (51–68)* | | 65 | | (60–70)* |
| Length, cm | 181 | (178–183) | 186 | (181–188)* | 176 | (171–182)* | 177 | (172–180)* | | 177 | | (174–182)* |
| Weight, kg | 80.5 | (74–88) | 82 | (78–87) | 82.5 | (76–92) | 84 | (77–92) | | 85 | | (80–95) |
| BSA, m^2^ | 2.0 | (1.9–2.1) | 2.0 | (2.0–2.1) | 2.0 | (1.9–2.2) | 2.0 | (1.9–2.1) | | 2.1 | | (1.9–2.2) |
| BMI, kg/m^2^ | 24 | (23–27) | 24 | (23–25) | 27 | (24–30)* | 27 | (25–29)* | | 27 | | (25–29)* |
| LVEF, % | 59 | (55–62) | 55 | (52–59)* | 26 | (20–34)* | 48 | (42–55)* | | 63 | | (59–68)* |
| LVEDV, ml | 194 | (172–215) | 252 | (225–273)* | 331 | (273–369)* | 181 | (157–204)* | | 171 | | (162–220) |
| LVESV, ml | 78 | (72–89) | 112 | (97–127)* | 243 | (177–281)* | 93 | (76–115)* | | 62 | | (56–92)* |
| LVSV, ml | 114 | (104–128) | 138 | (130–150)* | 81 | (65–96)* | 85 | (74–98)* | | 108 | | (89–124) |
| LVM, g | 123 | (112–136) | 160 | (144–179)* | 185 | (150–215)* | 133 | (117–147)* | | 138 | | (120–161)* |
| LVEDVI, ml/m^2^ | 96 | (89–106) | 123 | (117–132)* | 156 | (136–188)* | 90 | (80–102)* | | 84 | | (76–104)* |
| LVESVI, ml/m^2^ | 40 | (35–45) | 55 | (48–61)* | 114 | (89–147)* | 46 | (38–57)* | | 29 | | (25–43)* |
| LVSVI, ml/m^2^ | 57 | (51–64) | 69 | (65–72)* | 42 | (33–50)* | 43 | (37–49)* | | 51 | | (48–59)* |
| LVMI, ml/m^2^ | 62 | (57–69) | 79 | (72–87)* | 90 | (75–102)* | 67 | (58–73)* | | 68 | | (61–77) |

Characteristics for the whole validation/derivation cohort (*n=*537) shown as median and interquartile range where * denotes *p* < 0.05 compared to the healthy volunteers.. BSA=body surface area; CRT=cardiac resynchronization therapy-candidates (patients with heart failure); LVEDV(I)=left ventricular end-diastolic volume (indexed to BSA); LVEF=left ventricular ejection fraction; LVEDV(I)=left ventricular end-diastolic volume (indexed to BSA); LVESV(I)=end-systolic volume (indexed to BSA); LVM(I)=left ventricular mass(indexed to BSA); LVSV(I)=left ventricular stroke volume (indexed to BSA).

**Table S2a. Characteristics and results for the female subjects of the *mixed cohort*.**

| ***Females*** | **Healthy volunteers** | | **Fabry** | | **LVH** | |
| --- | --- | --- | --- | --- | --- | --- |
| Number, *n* | 35 |  | 86 |  | 60 |  |
| Age, years | 29 | (23–49) | 45 | (33–55)* | 61 | (47–68)* |
| Height, cm | 169 | (164–171) | 164 | (158–170)* | 166 | (162–170) |
| Weight, kg | 65 | (60–70) | 66 | (61–76) | 68 | (61–78) |
| BSA, m^2^ | 1.8 | (1.7–1.8) | 1.7 | (1.6–1.9) | 1.7 | (1.7–1.9) |
| BMI, kg/m^2^ | 23 | (21–25) | 24 | (22–28)* | 25 | (21–28) |
| LVEDV, ml | 152 | (131–169) | 120 | (107–134)* | 182 | (138–232)* |
| LVEDVI, ml/m^2^ | 88 | (77–95) | 68 | (61–76)* | 100 | (80–127)* |
| LVESV, ml | 60 | (52–70) | 29 | (22–35)* | 89 | (63–147)* |
| LVESVI, ml/m^2^ | 36 | (29–39) | 17 | (13–21)* | 49 | (35–78)* |
| LVSV, ml | 89 | (82–97) | 91 | (81–101) | 82 | (64–103)* |
| LVSVI, ml/m^2^ | 51 | (47–57) | 52 | (47–56) | 46 | (37–53)* |
| LVEF, % | 60 | (58–63) | 76 | (71–80)* | 46 | (35–61)* |
| LVM, g | 89 | (77–97) | 104 | (85–124)* | 149 | (134–175)* |
| LVMI, g/m^2^ | 51 | (45–56) | 56 | (49–71)* | 83 | (77–96)* |
| GT, mm | 6.0 | (5.6–6.3) | 7.5 | (6.4–9.0)* | 8.7 | (7.9–9.5)* |
| GTI, mm/m^2^ | 3.4 | (3.1–3.6) | 4.2 | (3.7–5.2)* | 4.8 | (4.3–5.5)* |

Only the groups that were changed from the derivation/validation cohort are shown. * denotes *p* < 0.05 compared to the healthy volunteers.

**Table S2b. Characteristics and results of the male subjects of the *mixed cohort*.**

| ***Males*** | **Healthy volunteers** | | **Fabry** | | **LVH** | |
| --- | --- | --- | --- | --- | --- | --- |
| Number, *n* | 64 |  | 58 |  | 103 |  |
| Age, years | 32 | (26–49) | 42 | (34–54)* | 59 | (46–69)* |
| Height, cm | 181 | (177–184) | 178 | (172–185)* | 180 | (174–184) |
| Weight, kg | 80 | (73–87) | 77 | (65–83)* | 87 | (78–100)* |
| BSA, m^2^ | 2.0 | (1.9–2.1) | 2.0 | (1.8–2.1)* | 2.1 | (1.9–2.2)* |
| BMI, kg/m^2^ | 24 | (23–27) | 23 | (20–26) | 27 | (24–31)* |
| LVEDV, ml | 194 | (172–215) | 150 | (133–175)* | 239 | (191–299)* |
| LVEDVI, ml/m^2^ | 96 | (90–107) | 80 | (68–90)* | 114 | (91–151)* |
| LVESV, ml | 77 | (71–89) | 39 | (31–55)* | 135 | (87–191)* |
| LVESVI, ml/m^2^ | 39 | (35–46) | 21 | (16–28)* | 63 | (41–97)* |
| LVSV, ml | 116 | (104–132) | 108 | (90–130) | 99 | (77–124)* |
| LVSVI, ml/m^2^ | 58 | (53–64) | 54 | (48–66) | 48 | (37–60)* |
| LVEF, % | 60 | (56–62) | 72 | (67–78)* | 44 | (30–58)* |
| LVM, g | 126 | (113–146) | 182 | (148–246)* | 221 | (195–243)* |
| LVMI, g/m^2^ | 63 | (58–70) | 93 | (72–127)* | 103 | (96–115)* |
| GT, mm | 7.1 | (6.8–7.6) | 10.8 | (8.7–14.3)* | 10.1 | (9.1–11.3)* |
| GTI, mm/m^2^ | 3.6 | (3.3–3.8) | 5.5 | (4.5–7.1)* | 4.9 | (4.3–5.5)* |

Only the groups that were changed from the derivation/validation cohort are shown. Characteristics for the healthy volunteers and the patient groups shown as median and interquartile range and ***** denotes *p* < 0.05 compared to healthy volunteers. BMI=body mass index; BSA=body surface area; CRT=cardiac resynchronization therapy-candidates (patients with heart failure); LVEDV(I)=left ventricular end-diastolic volume (indexed to BSA); LVESV(I)=left ventricular end-systolic volume (indexed to BSA); LVH=left ventricular hypertrophy; LVSV(I)=left ventricular stroke volume (indexed to BSA); LVEF=left ventricular ejection fraction; LVM(I)=left ventricular mass (indexed to BSA).

**Table S3a. Nominal characteristics for the patients of the *survival cohort*.**

|  | **Normal** | | **Concentric remodeling** | | **Eccentric hypertrophy** | | **Concentric hypertrophy** | | ***p*** |
| --- | --- | --- | --- | --- | --- | --- | --- | --- | --- |
| *n* | 1133 | | 189 | | 89 | | 164 | |  |
| Males | 651 | 57% | 112 | 59% | 58 | 65% | 90 | 55% | 0.43 |
| Death or HHF | 202 | 18% | 51 | 27% | 36 | 40% | 62 | 38% | <0.001 * |
| Death | 151 | 13% | 41 | 22% | 27 | 30% | 38 | 23% | <0.001 * |
| LGE by CMR | 381 | 34% | 90 | 48% | 63 | 71% | 107 | 65% | <0.001 * |
| Infarction by CMR | 216 | 19% | 42 | 22% | 33 | 37% | 43 | 26% | <0.001 * |
| Non-ischemic scar by CMR | 185 | 16% | 55 | 29% | 34 | 38% | 68 | 41% | <0.001 * |
| Diabetes mellitus type 2 | 183 | 16% | 70 | 37% | 15 | 17% | 54 | 33% | <0.001 * |
| Hypertension | 501 | 44% | 137 | 72% | 38 | 43% | 117 | 71% | <0.001 * |
| CABG prior to CMR | 77 | 7% | 23 | 12% | 8 | 9% | 14 | 9% | 0.07 |
| PCI prior to CMR | 138 | 12% | 31 | 16% | 14 | 16% | 19 | 12% | 0.31 |

The patients are characterized as being either normal (normal GT and LVMI), or having concentric remodeling (high GT, normal LVMI), eccentric hypertrophy (high LVMI, normal GT), or concentric hypertrophy (high LVMI and GT). Data shown as *n* and percentage and *p*-value calculated using the Fisher’s exact test for difference between the groups, and * denotes *p* < 0.05.

**Table S3b. Numerical characteristics for the patients of the *survival cohort*.**

|  | **Normal** | | **Concentric remodeling** | | **Eccentric hypertrophy** | | **Concentric hypertrophy** | | ***p*** |
| --- | --- | --- | --- | --- | --- | --- | --- | --- | --- |
| *n* | 1133 | | 189 | | 89 | | 164 | |  |
| Age at CMR, years | 56 | (44–65) | 60 | (52–68) | 57 | (46–65) | 57 | (47–66) | 0.001 * |
| BMI, kg/m^2^ | 28 | (24–33) | 35 | (29–41) | 26 | (23–30) | 30 | (25–36) | <0.001 * |
| BSA, m^2^ | 2.0 | (1.8–2.2) | 2.2 | (2.0–2.5) | 2.0 | (1.7–2.1) | 2.1 | (1.9–2.3) | <0.001 * |
| Height, m | 1.73 | (1.64–1.80) | 1.73 | (1.63–1.83) | 1.73 | (1.65–1.78) | 1.73 | (1.65–1.80) | 0.71 |
| Weight, kg | 84 | (72–98) | 103 | (85–122) | 80 | (64–93) | 90 | (77–109) | <0.001 * |
| LVEDVI, ml/m^2^ | 80 | (67–97) | 68 | (56–83) | 161 | (137–196) | 112 | (92–131) | <0.001 * |
| LVESVI, ml/m^2^ | 32 | (25–46) | 26 | (19–38) | 123 | (94–159) | 62 | (40–90) | <0.001 * |
| LVEF, % | 59 | (50–64) | 61 | (51–66) | 23 | (17–32) | 40 | (31–56) | <0.001 * |
| LVMI, g/m^2^ | 50 | (42–59) | 62 | (54–72) | 87 | (82–93) | 91 | (81–106) | <0.001 * |
| GT, mm | 6 | (6–7) | 9 | (8–9) | 7 | (7–8) | 9 | (9–10) | <0.001 * |
| GTI, mm/m^2^ | 3 | (3–4) | 4 | (4–4) | 4 | (3–4) | 4 | (4–5) | <0.001 * |
| eGFR, ml/min/1.73 m^2^ | 90 | (73–100) | 85 | (65–99) | 81 | (66–91) | 81 | (60–95) | <0.001 * |

The patients are characterized as being either normal (normal GT and LVMI), or having concentric remodeling (high GT, normal LVMI), eccentric hypertrophy (high LVMI, normal GT), or concentric hypertrophy (high LVMI and GT). Data shown as median (interquartile range) and *p*-value calculated using the Kruskall–Wallis test for difference between the four groups, * denotes *p* < 0.05. BMI=body mass index; BSA=body surface area; CABG=coronary artery bypass grafting; CMR=cardiovascular magnetic resonance; eGFR=estimated glomerular filtration rate, using the MDRD formula; HHF=hospitalization for heart failure; LGE=late gadolinium enhancement; LVEDVI=left ventricular end-diastolic volume indexed (to BSA); LVEF=left ventricular ejection fraction; LVMI=left ventricular mass indexed (to BSA); LVESVI=left ventricular end-systolic volume indexed (to BSA); GT(I)=global wall thickness (indexed to BSA); PCI=percutaneous coronary intervention.

**Table S4a. Nominal characteristics for the patients of the survival cohort with *normal findings*.**

|  | **All** | | **Normal GT** | | **High GT** | | ***p*** |
| --- | --- | --- | --- | --- | --- | --- | --- |
| *n* | 326 | | 300 | | 26 | |  |
| Female sex | 147 | (45 %) | 140 | (47 %) | 7 | (27 %) | 0.06 |
| Age | 47.5 | (33–59) | 47 | (32–59) | 56 | (40–62) | 0.04 * |
| Death or HHF | 29 | (9 %) | 21 | (7 %) | 8 | (31 %) | <0.001 * |
| Death | 21 | (6 %) | 15 | (5 %) | 6 | (23 %) | 0.003 * |
| LGE by CMR | 0 |  | 0 |  | 0 |  | - |
| Infarction by CMR | 0 |  | 0 |  | 0 |  | - |
| Non-ischemic scar by CMR | 0 |  | 0 |  | 0 |  | - |
| Diabetes mellitus type 2 | 27 | (8 %) | 22 | (7 %) | 5 | (19 %) | 0.05 |
| Hypertension | 106 | (33 %) | 91 | (30 %) | 15 | (58 %) | 0.008 * |
| CABG prior to CMR | 6 | (2 %) | 4 | (1 %) | 2 | (1 %) | 0.08 |
| PCI prior to CMR | 16 | (5 %) | 11 | (4 %) | 5 | (19 %) | 0.005 * |

Data is shown for the subgroup of the survival cohort with normal findings (no LGE, and findings within the normal range per sex for LVEDVI, LVMI, and LVEF). Data shown as *n* and percentage and *p*-value calculated using the Fisher’s exact test for difference between the groups, and * denotes *p* < 0.05.

**Table S4b. Numerical characteristics for the patients of the survival cohort with *normal findings*.**

|  | **Females** | | **Males** | | ***p*** |
| --- | --- | --- | --- | --- | --- |
| *n* | 147 | | 179 | |  |
| Age at CMR, years | 46 | (34–59) | 48 | (30–59) | 0.9 |
| BMI, kg/m^2^ | 27 | (23–33) | 28 | (25–32) | 0.05 |
| BSA, m^2^ | 1.8 | (1.7–2.0) | 2.1 | (2.0–2.3) | <0.001 * |
| Height, m | 1.65 | (1.60–1.70) | 1.80 | (1.75–1.85) | <0.001 * |
| Weight, kg | 72 | (64–88) | 91 | (82–103) | <0.001 * |
| LVEDVI, ml/m^2^ | 78 | (70–86) | 88 | (80–98) | <0.001 * |
| LVESVI, ml/m^2^ | 30 | (27–34) | 35 | (31–41) | <0.001 * |
| LVEF, % | 61 | (58–65) | 60 | (56–63) | <0.001 * |
| LVMI, g/m^2^ | 45 | (42–50) | 58 | (53–65) | <0.001 * |
| GT, mm | 5.8 | (5.4–6.4) | 7.2 | (6.5–7.7) | <0.001 * |
| GTI, mm/m^2^ | 3.2 | (2.9–3.5) | 3.3 | (3.0–3.7) | 0.002 * |
| eGFR, ml/min/1.73 m^2^ | 90 | (80–105) | 90 | (80–105) | 0.92 |

The patients are characterized as being either normal (normal GTI and LVMI), or having concentric remodeling (high GTI, normal LVMI), eccentric hypertrophy (high LVMI, normal GTI), or concentric hypertrophy (high LVMI and GTI). Data shown as median (interquartile range) and *p*-value calculated using the Kruskall–Wallis test for difference between the four groups, * denotes *p* < 0.05. BMI=body mass index; BSA=body surface area; CABG=coronary artery bypass grafting; CMR=cardiovascular magnetic resonance; eGFR=estimated glomerular filtration rate, using the MDRD formula; HHF=hospitalization for heart failure; LGE=late gadolinium enhancement; LVEDVI=left ventricular end-diastolic volume indexed (to BSA); LVEF=left ventricular ejection fraction; LVMI=left ventricular mass indexed (to BSA); LVESVI=left ventricular end-systolic volume indexed (to BSA); GT(I)=global wall thickness (indexed to BSA); PCI=percutaneous coronary intervention.
