## Supplementary figures and images for "Left ventricular mass and global wall thickness – prognostic utility and characterization of left ventricular hypertrophy"

### Supplementary Figure 1

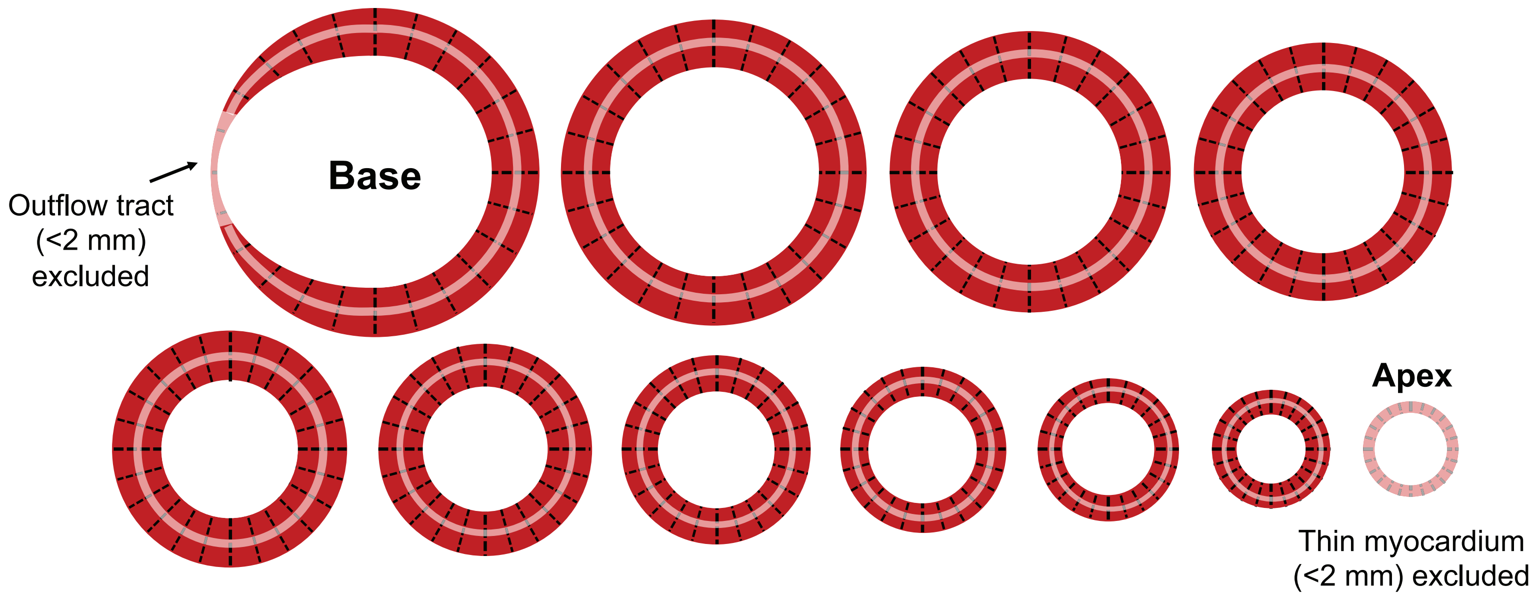

### Supplementary Figure 1

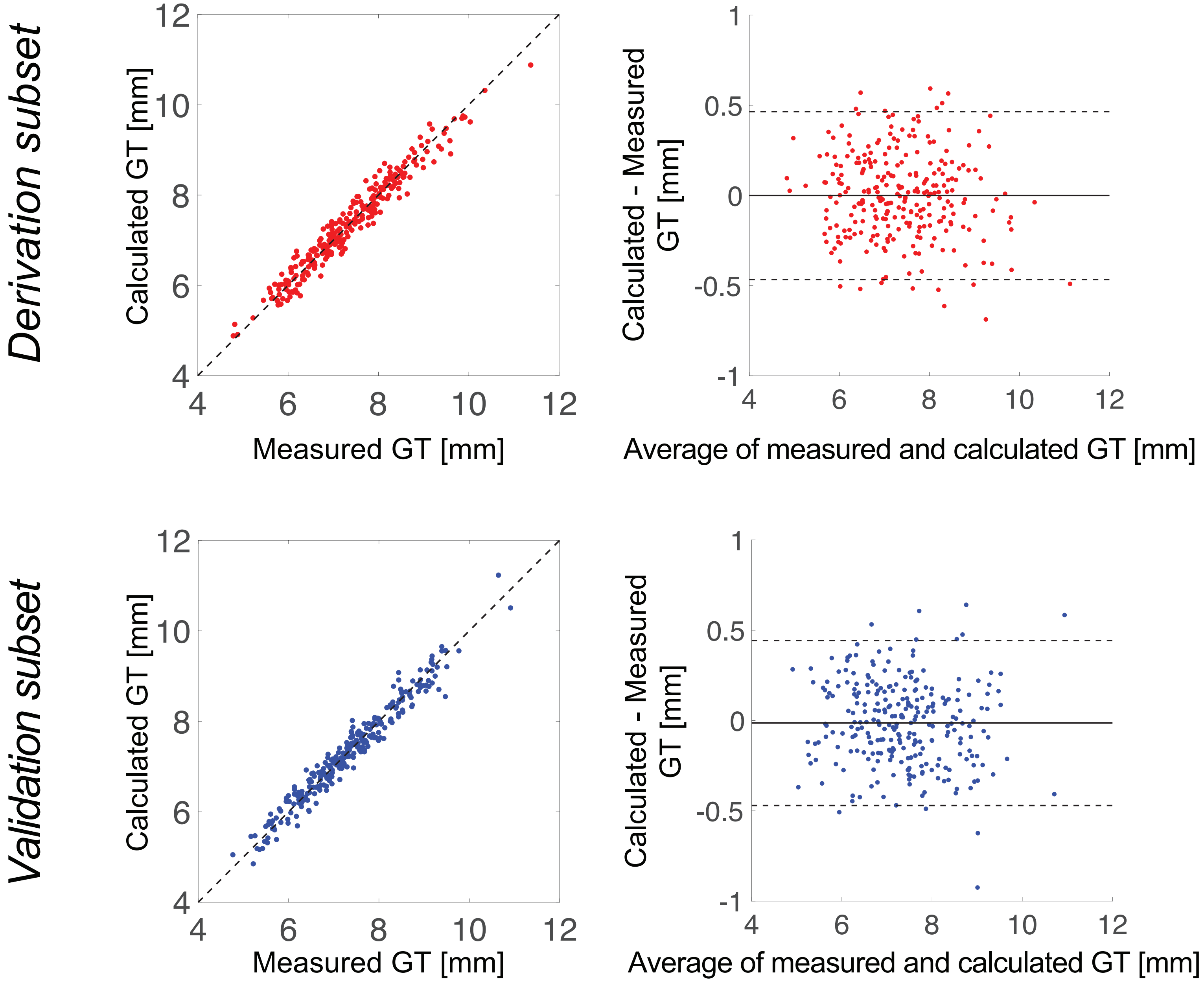
